# BrainTRACE (Brain Tumor Registration and Cortical Electrocorticography): A Novel Tool for Localizing Electrocorticography Electrodes in Patients with Brain Tumors

**DOI:** 10.1101/2025.05.07.25327167

**Authors:** Sena Oten, Sanjeev Herr, Vardhaan Ambati, Youssef Sibih, Katie Lu, Jasleen Kaur, Shawn L. Hervey-Jumper, David Brang

**Author notes:** Authors contributed equally (Shawn L. Hervey-Jumper, David Brang).

## Abstract

**Background:** Intraoperative electrocorticography (ECoG) plays a critical role in clinical care and neuroscience research, enabling precise mapping of functional cortex. However, localizing subdural electrodes in patients with brain tumors presents unique challenges due to altered neuroanatomy and the impracticality of acquiring extraoperative computed tomography (CT). To address this gap, we developed BrainTRACE, a novel MATLAB tool that combines magnetic resonance imaging (MRI), cortical vascular reconstructions, and intraoperative photography for accurate subdural electrode grid placement.

**Methods:** Preoperative MRI, cortical photography, and subdural electrode array data were recorded from patients with diffuse glioma and brain metastasis. BrainTRACE generated three-dimensional cortical surfaces, integrated vascular reconstructions, and enabled precise placement of electrode grids. Each electrode was placed based on cortical anatomy and vascular landmarks informed by intraoperative photographs. Novice and expert-level proficiency were quantified.

**Results:** Expert users achieved high consistency and accuracy, with an intraclass correlation coefficient (ICC) of 0.934 and a mean deviation of 4.3 mm from consensus placements. Novice users demonstrated lower reliability (ICC = 0.399) and greater variability, averaging a 16.3 mm deviation from consensus. These findings highlight the non-trivial nature of intraoperative ECoG localization, which requires neuroanatomical expertise for successful grid placement.

**Conclusion:** BrainTRACE enables accurate localization of intraoperative ECoG electrodes in brain tumor patients. By integrating anatomical images, intraoperative photographs, and vascular mapping, the tool addresses challenges posed by tumor-induced artifacts. While requiring that users have cortical neuroanatomical expertise, BrainTRACE provides a practical tool for neurosurgical and neuroscience applications, including brain malignancy, epilepsy, and deep brain stimulation procedures. BrainTRACE is freely available to researchers (https://github.com/dbrang/BrainTRACE).

**Highlights:** BrainTRACE addresses the challenge of ECoG electrode placement without the use of extra-operative imaging for clinical populations such as brain tumor patients

- A combined approach using preoperative MRI, cortical vascular reconstructions, and intraoperative photography ensures precise grid placement.
- BrainTRACE is freely available for research applications, offering a reproducible framework for accurate electrode placement.

## 1. Introduction

Electrocorticography (ECoG) is an electrophysiological monitoring technique in which subdural electrode arrays are placed along the brain’s cortical surface during neurosurgical procedures. Subdural arrays are used to record electrical signals generated by local populations of neurons, offering spatial resolution that is not present using non-invasive techniques such as electroencephalography (EEG) (Klaes, 2018). Historically, ECoG has been used predominantly in patients with epilepsy to localize epileptogenic zones and to electrically stimulate regions of the brain to minimize clinical deficits following tissue resection (Spencer et al., 1990; Engel et al., 2005). In epilepsy, electrodes are typically implanted chronically, with neuronal activity recorded in the extraoperative setting for one to two weeks to detect and localize a seizure’s anatomical onset or functional cortical regions (Moon et al., 2024).

Increasingly, ECoG is recorded in intraoperative contexts in multiple patient populations, including patients with brain tumors (Chang et al., 2011; Hervey-Jumper et al., 2015), Parkinson’s disease (Merk et al. 2022), cortical dysplasia (Sacino et al., 2016), cavernous malformation, and in some cases epilepsy. In patients with intrinsic brain tumors, ECoG is clinically used to localize the sensorimotor cortex and identify cortical regions essential for cognitive processes. Moreover, subdural array data recorded under clinical context has shed light on the extent to which neuronal populations are remodeled by cancer infiltration (Aabedi et al., 2021; Krishna et al., 2023).

Electrophysiological recordings performed under clinical context support basic research on cortical sensory and cognitive processes. For example, increased high gamma power (an index of population spiking rates; Ray et al., 2008) is observed in the fusiform gyrus in response to seeing images of faces (Ghuman et al., 2014), in the superior temporal gyrus in response to hearing sounds (Crone, Boatman, Gordon, & Hao, 2001; Ganesan et al. 2021), and pre-motor frontal regions during speech production (Kingyon et al., 2015). ECoG additionally holds significance in the realm of brain-computer interfaces (Awuah et al., 2024; Chang, 2015), where accurate mapping of brain function can aid in the development of assistive technologies for individuals with motor or communication impairments. The success of these applications heavily depends on the precise localization of ECoG electrodes, underscoring the need for reliable methods to achieve this goal.

Traditional methods for ECoG electrode localization were developed for the extraoperative examination of patients with epilepsy. In this situation, electrodes are localized using a postoperative computed tomography (CT) scan registered to a patient’s preoperative MRI. As metallic objects show bright white artifacts on a CT, this technique enables the accurate identification of individual electrode contacts. Several methods have been developed to facilitate this process (Branco et al., 2018; Gupta et al., 2014; Pieters et al., 2013) and include methods to counteract post-operative brain drift that can occur from chronic implantations (e.g., Brang et al., 2016). However, CT scans are rarely acquired for patients who receive intraoperative ECoG because the electrodes are not implanted chronically, and obtaining a CT would lengthen the duration of the surgery, adding additional risk to the patient, without clinical necessity. As a result, intraoperative ECoG localization requires alternative approaches.

While no standard approaches or software exist specifically for intraoperative ECoG registration, it is common to localize individual contacts based on cortical anatomy defined by intraoperative photographs taken with and without the grids implanted (Wellmer et al., 2002; Dalal et al., 2008). In this process, a skilled operator will identify the location of individual electrode contacts relative to gyral anatomy, providing a general atlas of electrode labels (e.g., contacts 1-3 on the grid were along the motor cortex). This process is time-consuming, requires substantial expertise, and does not allow fine-grained examination of anatomical positions or the projection of anatomical coordinates into a common space (e.g., MNI or Talairach). Alternatively, grids can be drawn onto 2D images of template brains, providing a rough estimation of their locations but with unknown accuracy.

Accordingly, there is a pressing need for an ECoG localization technique that does not require a postoperative CT and that is appropriate for patients with altered cortical anatomy, such as patients with a brain tumor. Therefore, we developed a tool that utilizes preoperative MRI scans, vascular reconstructions, intraoperative images, and neuro-navigation tools with a custom MATLAB graphical user interface (GUI) to localize electrode grids on patients who have undergone awake craniotomies for tumor resection (**Figure 1**). BrainTRACE (Brain Tumor Registration and Cortical Electrocorticography): addresses the unique challenges of localizing ECoG electrodes in patients with brain tumors, allowing clinicians to align and place electrodes, enhancing the accuracy of electrode localization, and is freely available to researchers (https://github.com/dbrang/BrainTRACE).

**Figure 1.**
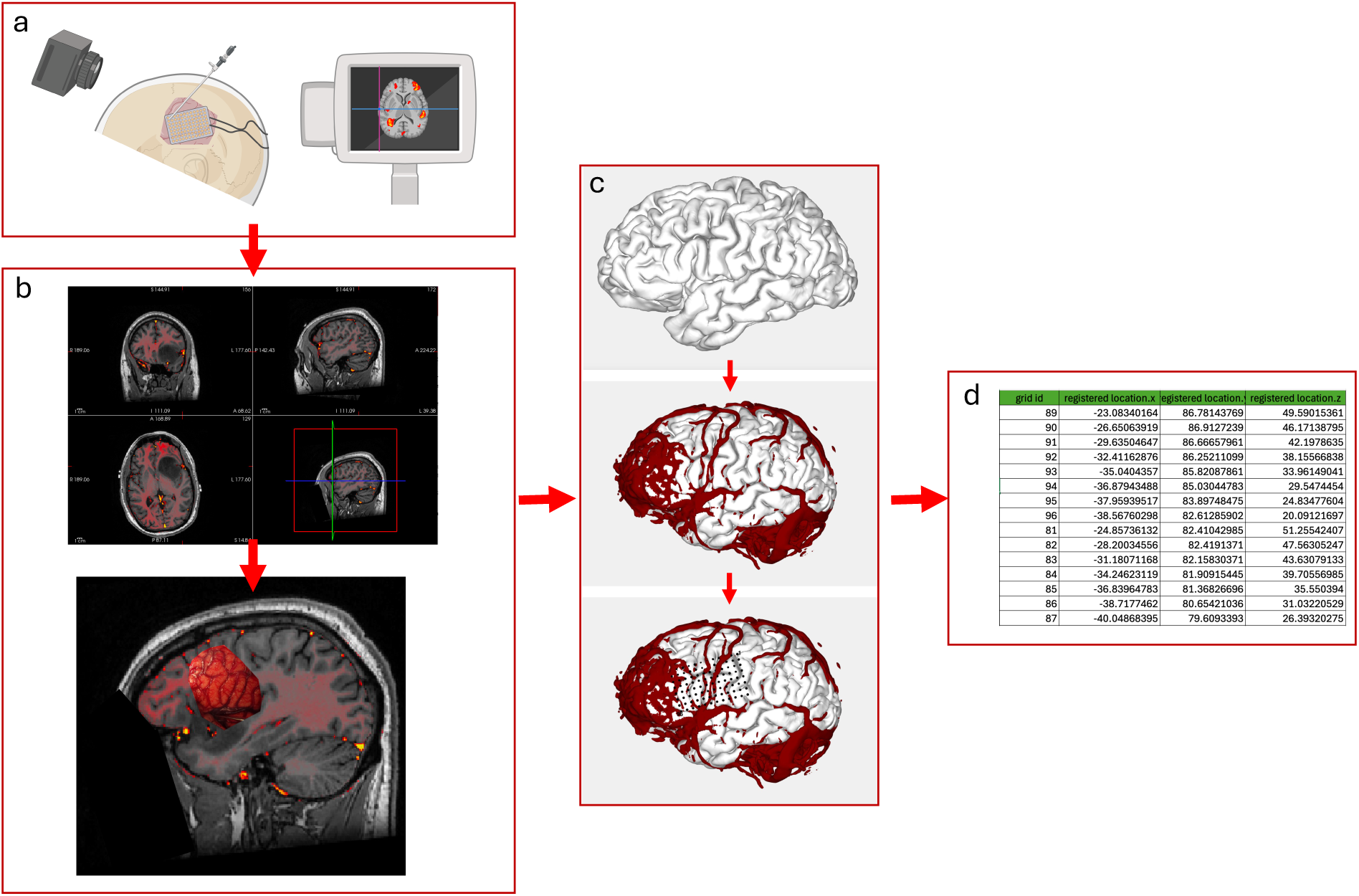
Workflow for electrode grid placement and coordinate extraction using the BrainTRACE software. **(a)** Intraoperative photographs are captured with and without the electrode grid placed on the brain. Neuronavigational tools can optionally be used to localize the grid corners. **(b)** Freeview or other MRI viewing software is utilized to visualize the tumor and associated cortical vasculature. **(c)** BrainTRACE enables visualization of the 3D reconstruction of the brain and vasculature, facilitating precise localization of the electrode grid within the anatomical space. **(d)** The XYZ coordinates of each electrode are exported for downstream analysis and integration into clinical or research workflows.

## 2. Methods

Patients were consented through IRB-17-23215 at the University of California, San Francisco. The IRB of the University of California, San Francisco gave ethical approval for this work. Deidentified Digital Imaging and Communications in Medicine (DICOM) images were obtained using Automated Image Retrieval (AIR) from UCSF’s picture archiving and communication system (PACS). Preoperative T1 MRI and post-gadolinium contrast T1 scans from each patient were used to create three-dimensional cortical and vascular brain reconstructions. Using the antsBrainExtraction.sh script from Advanced Normalization Tools (ANTs) (Avants et al., 2011), a brain extraction was performed to extract the skull and dura to create a clean image of the isolated brain. Before pre-processing these files, the quality of the extraction and mask were reviewed using FreeSurfer’s Freeview tool (Fischl 2012).

Our protocol used 20-contact (4×5) electrode grids with 10 mm spacings and/or 96-contact (8×12) with 5 mm spacing on patients undergoing awake brain-mapping craniotomies for tumor resection. Electrodes were secured under the dura to prevent shifting during the recordings. Intraoperative images were taken with and without the grid secured onto the cortex. Intraoperative photos and neuro-navigation coordinates of the corner of each grid were used to anatomically pinpoint the electrode contacts on the cortical reconstruction in the BrainTRACE GUI. All data produced in the present study are available upon reasonable request to the authors

### 2.1 Pre-Processing

BrainTRACE requires only an anatomical image of the brain extracted from the skull to function. We recommend using individual patient anatomy, but an MNI brain or other templates may be used. The software accepts .mgz and .pial file types as output from the Freesurfer recon-all pipeline. We recommend two processes to generate this image. First, the recon-all-clinical software (Billot et al., 2023) can be used to generate a cortical reconstruction for each hemisphere while filling in lesions and cortical abnormalities. It is recommended to create a reconstruction of the vasculature using a post-gadolinium T1 MRI image. Optional scripts have been provided on GitHub along with the BrainTRACE software to complete this process using Statistical Parametric Mapping (SPM) (Ashburner and Friston, 2005).

### 2.2 Using the BrainTRACE Software

A complete guide is available at protocols.io and additional information on the software on github: https://www.protocols.io/private/DB6CE588004C11F09F880A58A9FEAC02

https://github.com/dbrang/BrainTRACE

#### Step 1: Loading brain extracted volume or reconstructed cortical surface (Fig. 1a)

To load a brain extracted volume or surface into the software, click the “Select Brain File” button and select the corresponding .mgz or .pial surface file. If choosing a .pial surface, the software will then prompt the user to select a corresponding .mgz volumetric file to extract RAS transformation information that ensures alignment between the MRI, coordinates, and surfaces. Tumor-burdened brains have differing amounts of success with brain reconstructions (Fig. 3). The GUI will then show the user its estimate of the isovalue threshold generated from the histogram of the data. BrainTRACE identifies a pixel value and includes all voxels with a pixel value above that threshold. The software typically identifies an acceptable threshold value, but users may adjust the threshold for optimal visualization by right-clicking on the file within “select elements to display” and selecting “threshold MGZ file.”

#### Step 2: Creation of the dura surface

BrainTRACE generates an estimate of the dura surface as the surface boundary of the brain. ECoG electrodes are then placed in relation to the dura as opposed to the brain’s surface. This is necessary as electrodes may otherwise fall into sulcal gaps. The dura is updated once the brain reconstruction image is adjusted for an optimal view of the sulcal anatomy. The GUI will prompt users to create dura automatically after loading a reconstruction or users may load an externally created dura surface. Alternatively, this step may be completed by right-clicking on the brain file within “select elements to display” and selecting “create dura.” The software will prompt the user for a dura shrink factor ranging from 0 to 1 and corresponding to the ‘shrink factor’ of the MATLAB ‘boundary’ function. The software computes the value automatically, and thus, the user does not need to adjust this value. Once users no longer need to view the dura, they may hide it by deselecting the dura element.

#### Step 3: Loading vasculature or additional surface information (Fig. 2b)

To load the reconstructed vasculature, click “load additional file” and select the post-gadolinium brain mask vasculature .mgz file. The GUI will once again show the user an estimate of the ideal isovalue threshold. In general, the user should adjust this value for optimal anatomically accurate modeling and reduction of artifacts. This will become useful when matching anatomical locations between the intraoperative photos and the brain reconstruction (Fig. 7). Once the threshold value is selected, the GUI will prompt the user for a transparency value. This is an opacity value between 0 and 1, with zero being completely transparent and one completely opaque.

**Figure 2.**
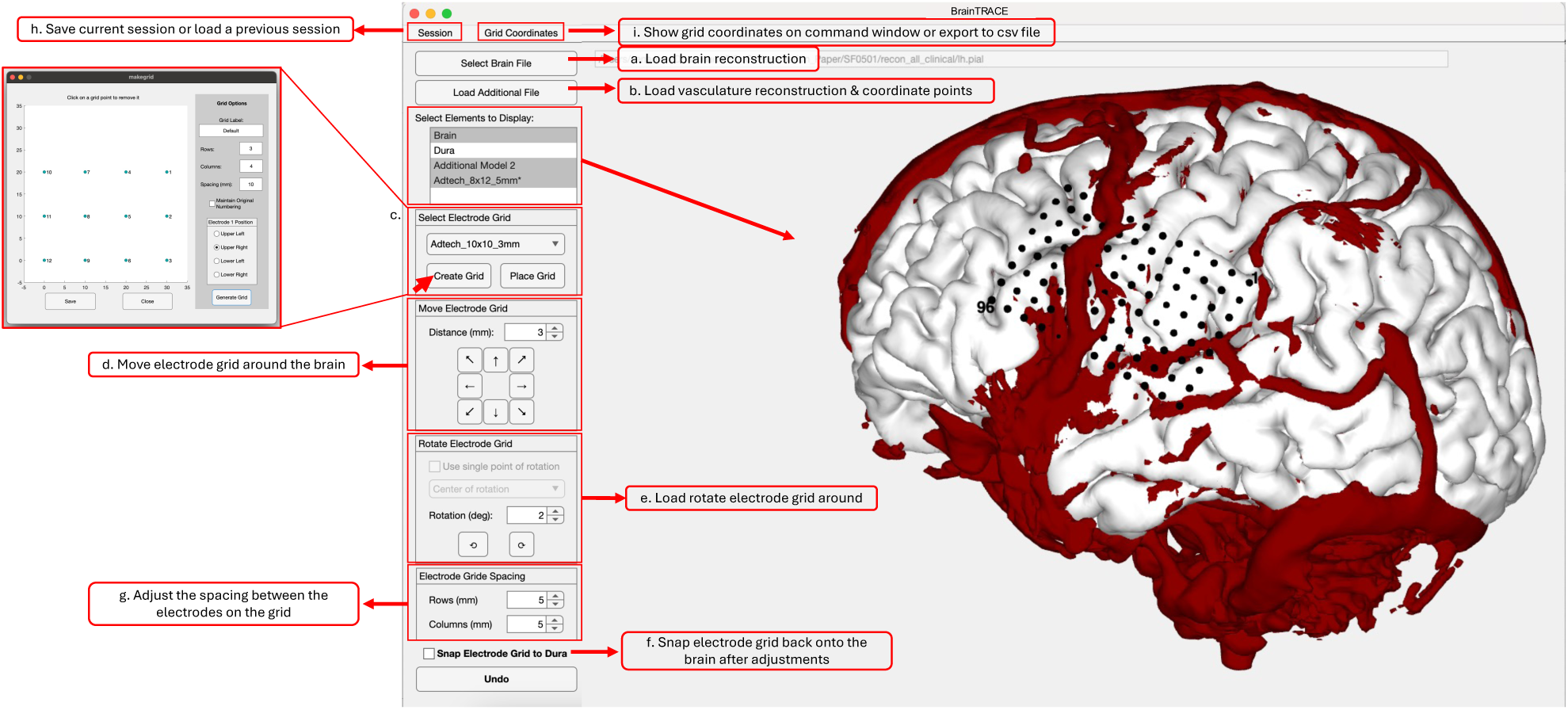
Graphical User Interface (GUI) of the BrainTRACE Software. **(a)** Interface for loading brain volumes or cortical surfaces, with tools for threshold adjustment and anatomical visualization. **(b)** Addition of vasculature and neuro-navigation coordinates, with transparency and alignment tools. **(c)** Electrode grid creation and placement, including default and custom grids, with adjustment options. **(d-f)** Tools for fine-tuning grid position, rotation, spacing, and snapping to account for anatomical changes. **(g)** Manage sessions and coordinate export for integration with external software or ECoG data analysis.

#### Step 4 (Optional): Using neuronavigation coordinates

To create circular points where the neuro-navigation coordinates were taken, users select “load additional file” (Fig. 2b) and select a file with CSV coordinates in an X, Y, or Z format. Coordinates can be helpful if neuronavigation coordinates are available, but users must be cautious that the coordinates are in the same anatomical space as the MRI. If the coordinates are in a different anatomical space, the brain can align using Procrustes transformation. A supplemental script is provided to support this function.

#### Step 5: Grid creation and placement (Fig. 2c)

The user selects which electrode array configuration they will use or create a new grid in the “select electrode grid” section of the GUI. The software comes pre-loaded with four default grids from Adtech, including (1) a 20 contact, 4×5, 10 mm spaced grid, (2) a 96 contact, 8×12, 5 mm spaced grid, (3) a 100 contact, 10×10, 3 mm spaced grid, and (4) a 8×1, 10 mm spaced strip. If the user creates a new grid configuration, they must add the number of electrodes in each row and column and the millimeter spacing between electrodes. Next, the user selects the corner to position electrode 1. Users can click on a grid point for removal if the array has missing points or the electrode grid is not rectangular. Generated grids are saved in the user’s Documents directory and can be copied to new users.

We assume that electrodes are a fixed distance (e.g., 10 mm center-to-center spacing standard for clinical applications). While these distances are physically set by the manufacturer, distances may vary slightly when placed along the cortical surface. This is due to imperfections in the reconstruction of the pial surface, intraoperative swelling of the brain, or protrusion of the brain due to the tumor, each of which may cause the actual geodesic distance of electrodes to differ slightly from the distances along the cortical surface. When this is the case, users should utilize the “electrode grid spacings” (Fig. 2g) section of the GUI interface. With this, users can adjust the spacing of the electrodes in a case-by-case basis.

To add the grid on top of the cortical reconstruction, users should click “place grid.” The GUI will then prompt the user to place the grid based on coordinates or manually select a point along the surface. If neural navigation coordinates are available and verified to match the anatomical space of the brain extraction, these may be inputted as XYZ coordinate values on the coordinate list. If the manual approach is chosen, the GUI will instruct the user to select a point where the grid should be placed on the model. The grid placement should be selected from visual anatomic or vascular correspondence to the intraoperative photos. Once the grid had been placed, millimetric adjustments should be made using the “move electrode grid” (Fig. 2d) controls. This will place the electrode grid at the approximate position but will not immediately find the vertices on the brain closest to each electrode. While making coordinate adjustments, the grid may lose some of the cardinal axes, and some electrode contacts may fall into the brain. Users should use the “rotate electrode grid” (Fig. 2e) function if the grid needs to be rotated. Users may rotate the grid at the center of the coordinates or pick a single point of rotation for the grid along. The latter is useful when one point on the electrode grid has been placed anatomically in the correct location and should not be moved. Once the electrode grid is placed at a close approximation to the intraoperative image, users may select the option to snap the electrode to the dura (Fig. 2f), which will recalculate the distances of all electrodes to the nearest placement on the dura.

Additional grids can be added in the same way as the initial grid. Grid names can be relabeled on the display elements by right-clicking. Grids may be recolored using this feature. Users need to define which grid they are trying to manipulate on the localization. This can be done by right-clicking on a grid and marking it as the active grid. The current active grid will have an asterisk (*) by the name on the display elements box.

#### Step 5: Saving/Loading Sessions and Exporting Coordinates

To save each session, users may select “Session” (Fig. 2h) and save as .mat file. To find the XYZ coordinates of each contact, users can go back to the command window on MATLAB, which automatically generates coordinates for each contact. On the GUI, users may select to export grid coordinates by clicking “Grid Coordinates” (Fig. 2i), which will save the coordinates as a .csv file. Users can choose a load session on the GUI to load and back up a previously saved session. Outputted coordinates can be loaded into programs for plotting data in a distributed format. Additionally, coordinates may be loaded into Freesurfer as a set of dot/data points for correlation with relative cortical anatomy.

### 2.3 Evaluation of the tool

The accuracy of subdural electrode placement using BrainTRACE was evaluated within and across users. Inter-rater reliability was analyzed by comparing grid placements among multiple users (*n*=4 novices including an undergraduate student and three medical students; *n*=3 experts including a research technician and two faculty) against a consensus placement; the consensus was established by the three expert users after completion of their individual placements. Intraclass correlation (ICC) was calculated to quantify the degree of agreement among users, providing a robust measure of consistency. Additionally, the time required for users to complete grid placements was recorded by novice users, and approximated by experts.

## 3. Results

Performance of BrainTRACE with different brain extraction and reconstruction tools was assessed using FreeSurfer’s recon-all, FreeSurfer’s recon-all-clinical, and ANTsBrainExtraction (Fig. 3). Overall, FreeSurfer’s recon-all-clinical tool consistently performed the best with tumor burdened brain MRI files, producing the least tumor artifact and the most defined sulci.

**Figure 3.**
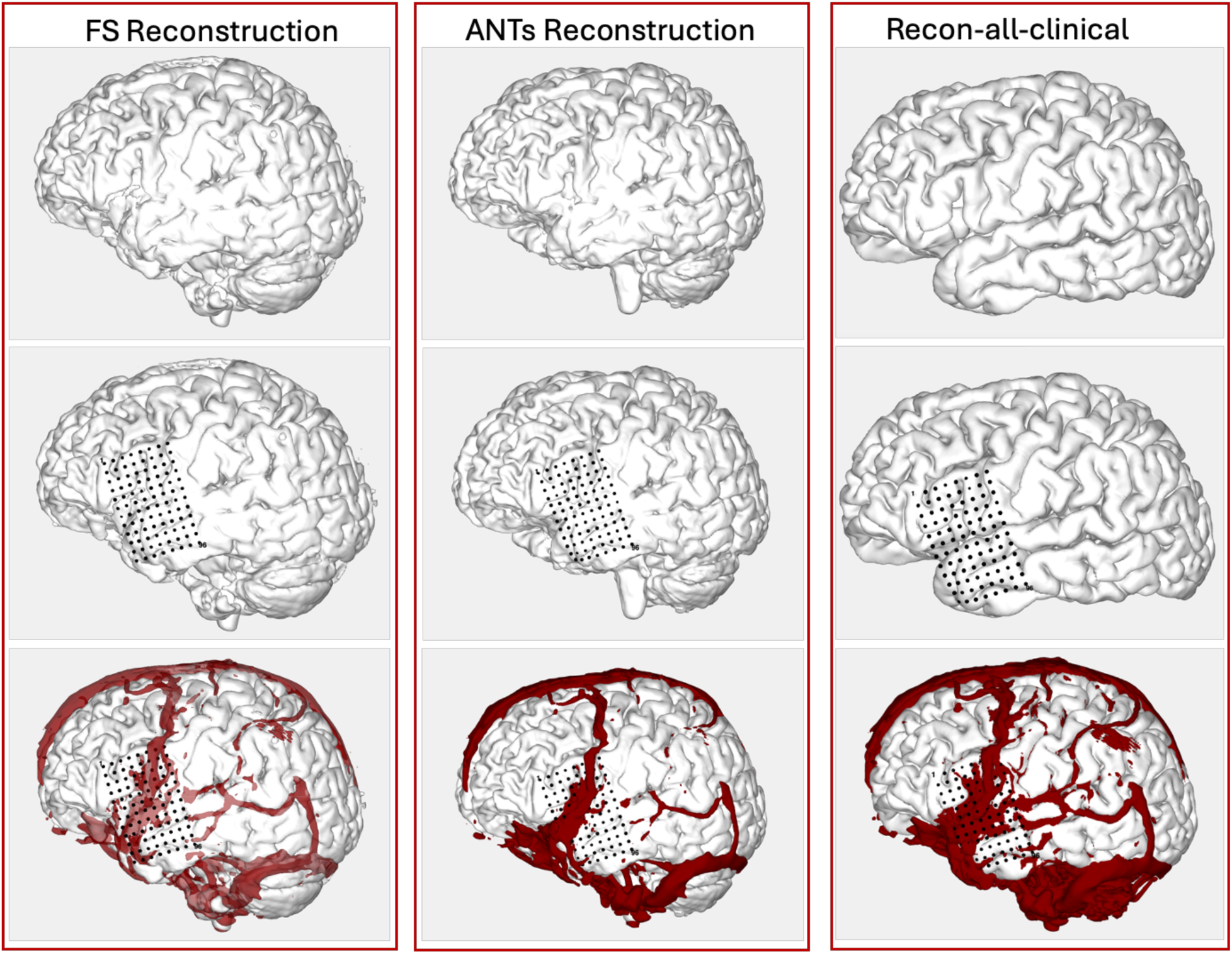
Performance of BrainTRACE software with different reconstruction pipelines. The first row presents a left-sided pial surface reconstruction. The second row adds a 96-channel grid to the left-sided pial surface reconstruction. The third row incorporates a vasculature reconstruction alongside the pial surface and 96-channel grid. Reconstructions using the Recon-all-clinical tool demonstrate higher detail in surface anatomy and reduced artifacts compared to alternative methods.

### Time-to-Completion

Registration times were measured across all users, with experts completing the process on average faster than novices (*p*<.001). Expert users required a mean registration time of 28.8 minutes per grid, whereas novices averaged 59.2 minutes, indicating that experience improves accuracy and efficiency. Novice users saw a significant (*p*<.001) reduction in the time it took to complete a grid placement throughout the 10 assigned registrations for this study (Fig. 4b). The final grid placement for each novice operator took 31 minutes (Range: 25-40) similar in time to the experts (mean = 21.3, range: 20-24).

**Figure 4.**
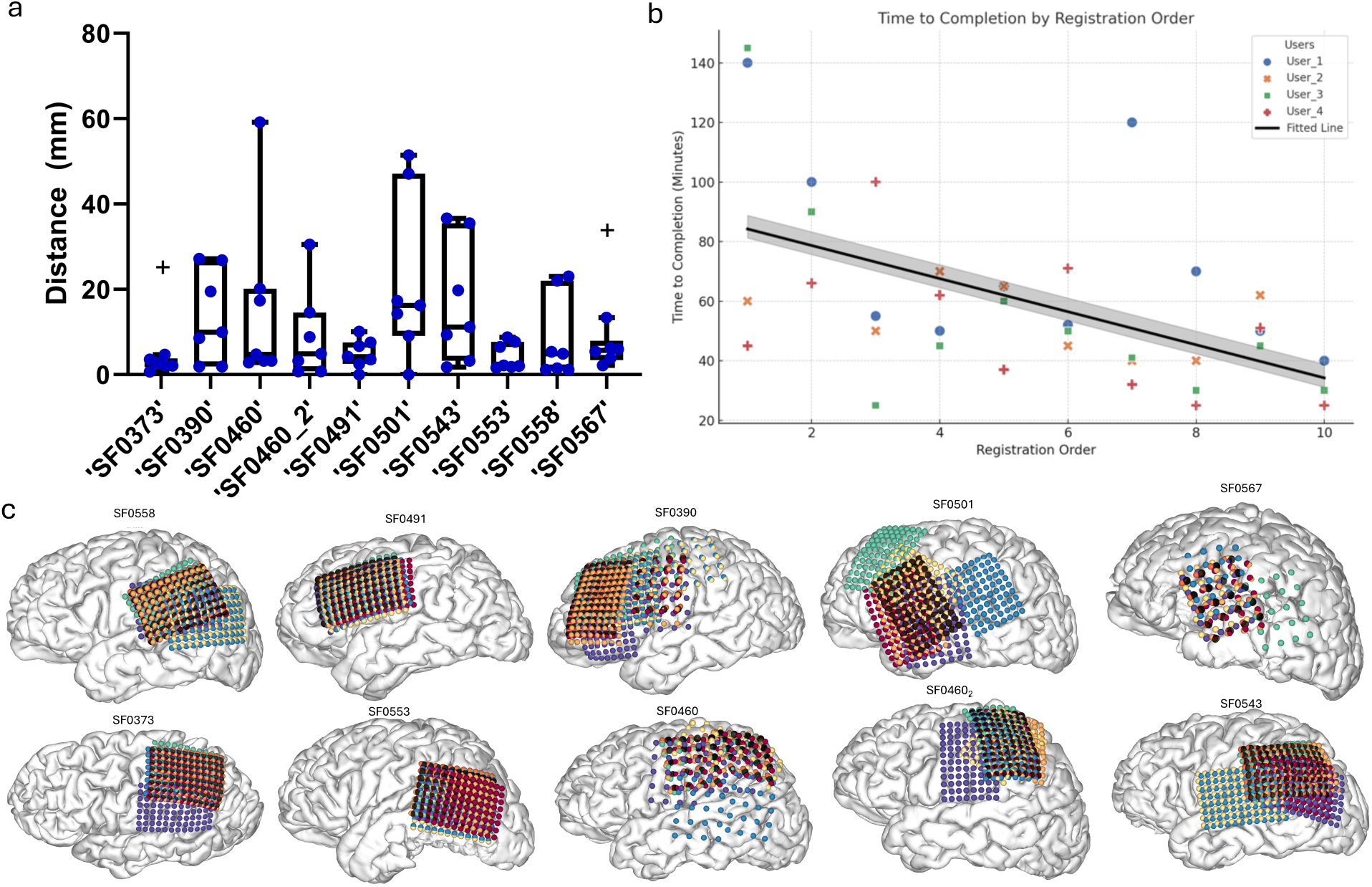
Variability and Speed Improvement in Grid Placement Performance Across Brains. **(a)** Average error relative to consensus placement across 10 patients, highlighting variability due to inter-brain differences. **(b)** Depiction of the improvement in grid placement speed over time in novice users, with a significant negative correlation (r² = 0.34, p < 0.001). **(c)** A visual representation of all user placements over the pial reconstructions of 10 patients. This graphic highlights inter-rater variability in grid placements and the consistency of this variability across patients. The black grid represents the consensus placement determined by three expert users, while colored grids denote individual user placements.

### Evaluation of Registration Accuracy and Inter-Rater Reliability

The accuracy and reliability of BrainTRACE were assessed by comparing electrode placements across patient 10 registrations performed by individuals with varying levels of expertise. Variability in grid placement accuracy was observed between brains, with some cases presenting greater challenges for precise electrode placement due to their unique neuroanatomical features and limited size of craniotomy windows (Fig. 4a, c). To compute the reliability of BrainTRACE across users, the study included two cohorts: experts (*n*=3), defined as individuals with formal neuroanatomy training and prior electrode placement experience, and novices (*n*=4) with minimal relevant neuroanatomy exposure. Accuracy was quantified as the distance from each operator’s grid placement relative to the consensus placement (mean Euclidean distance between each electrode). Experts achieved a mean distance of 4.3 mm, while novices displayed significantly larger variability, with a mean distance of 16.3 mm (Fig. 5a). Intraclass Correlation Coefficients (ICCs) further quantified inter-rater reliability, with experts achieving a high ICC of 0.934, indicating strong consistency in their placements. In contrast, novices exhibited considerably lower reliability, with an ICC of 0.399, underscoring the challenges faced by inexperienced users in achieving consistent and accurate placements (Fig. 5b, c). These findings emphasize the importance of neuroanatomical training for optimal electrode placement.

### Grid Placement Accuracy Between Experts and Novices

**Figure 5.**
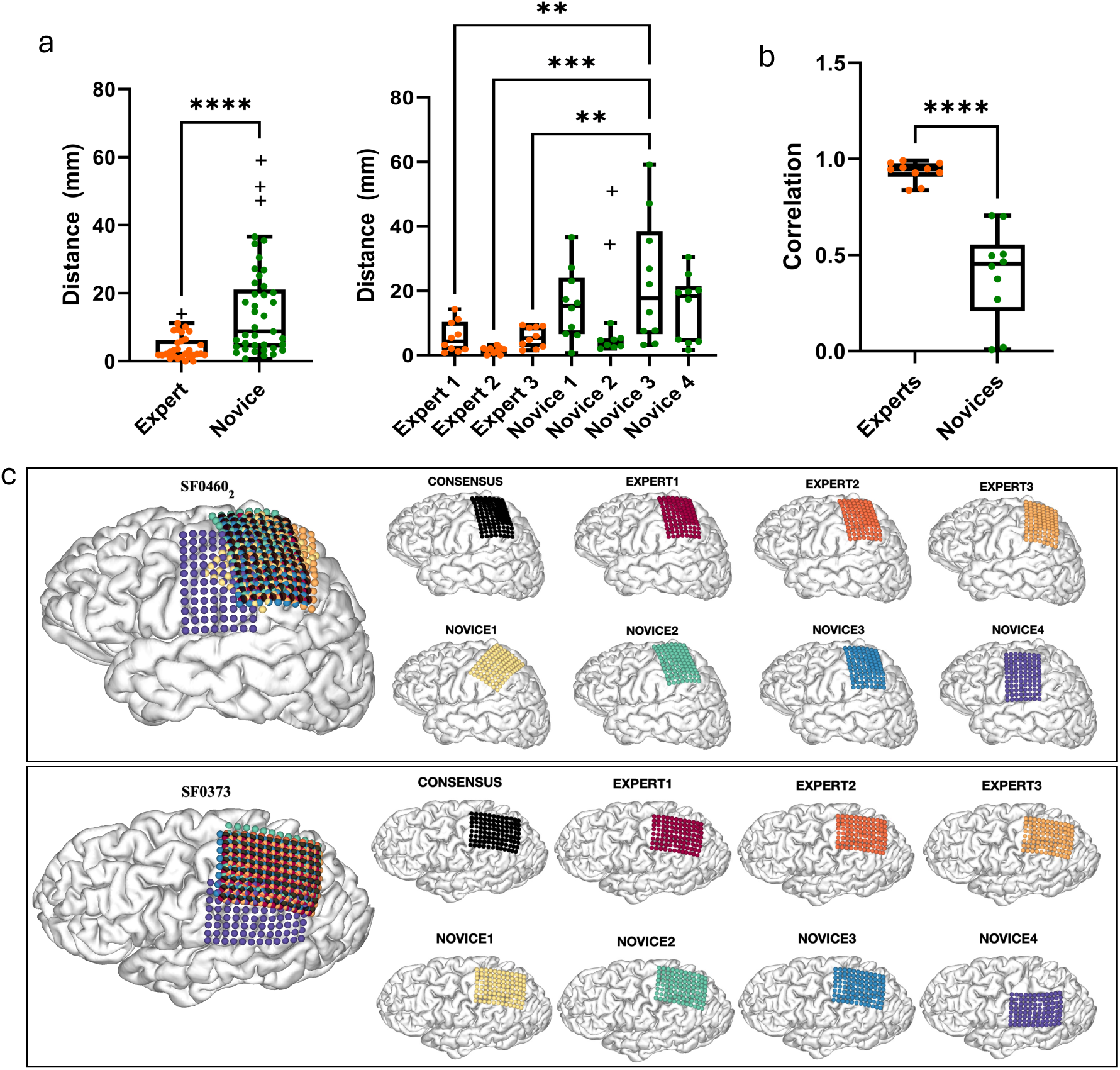
Assessing Consistency and Variability in Grid Placement Across Expert and Novice Users. **(a)** Comparison of the distance from consensus placements between expert and novice users. The first graph aggregates data from all experts and novices, demonstrating a significantly lower distance from the consensus for experts compared to novices (p < 0.001). The second graph provides a detailed breakdown of individual novice and expert performances. **(b)** Intraclass correlation (ICC) values for grid placements by experts and novices. Experts exhibited significantly higher ICC, indicating greater consistency (p < 0.001). **(c)** Examples of grid placement accuracy and consistency for two patients (SF0460 and SF0373) using the BrainTRACE electrode registration software. Patient SF0460 demonstrated increased variability in user placements compared to SF0373, with more pronounced differences among novice users. Across both patients, novices exhibited higher variability in grid placement compared to experts, relative to the consensus. Different colors represent placements by individual users, while the consensus placement is depicted in black for reference.

### Incorporation of Image Alignment Tools

In several cases, BrainTRACE’s image alignment add-on tool proved to be beneficial in refining electrode placement (Fig. 6). Specifically, the intraoperative photos provided limited information or ambiguous landmarks in some cases. In such instances, overlaying images of the brain with and without the grid that were aligned and warped was reported by several users to be effective in making the registration process more manageable. These supplemental tools are available: https://github.com/dbrang/IntraOp_Image_Registration_Tools.

**Figure 6.**
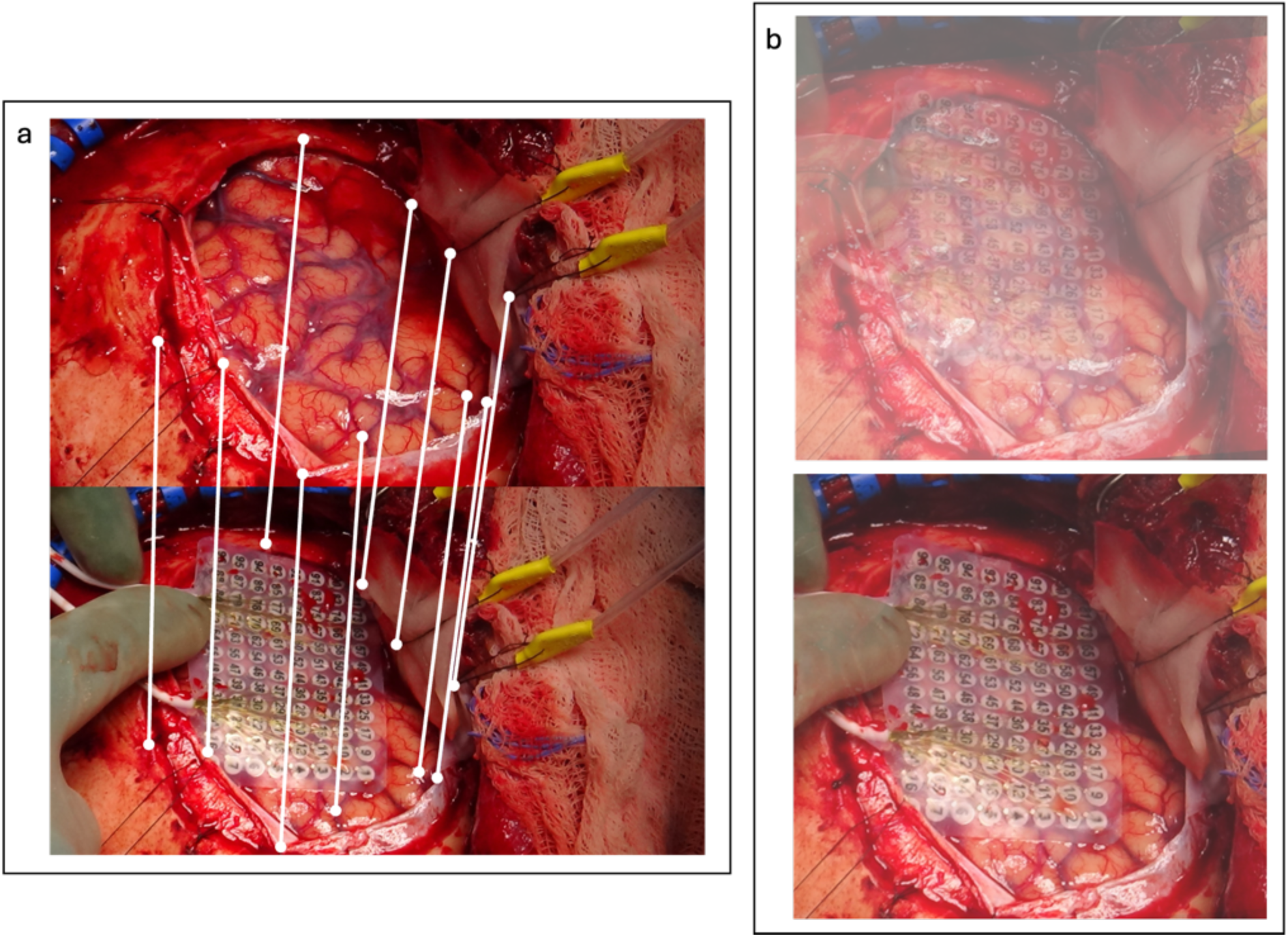
Example of intraoperative photos aligned to account for differences in photo angles to match sulcal anatomy and vasculature. **(a)** Shows the manually matched points between two images using the add on alignment tool. **(b)** Illustrates different transparency levels on the overlayed grid and no-grid photos to aid in matching electrodes to anatomical location

**Figure 7.**
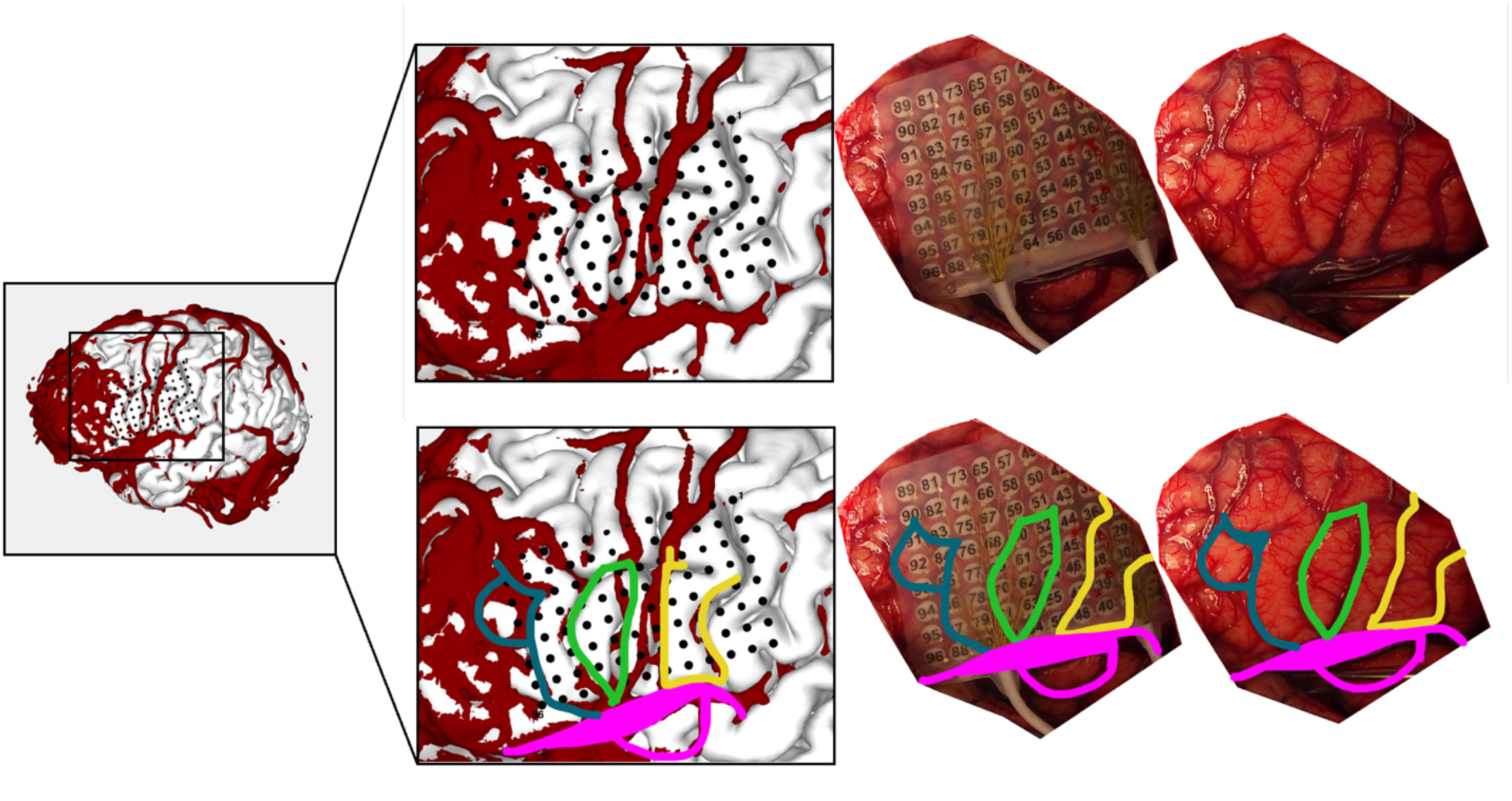
Overview of BrainTRACE functionality for matching sulcal and vascular anatomy between intraoperative photographs and 3D reconstructions in an illustrative case. This example highlights a successful electrode registration for a patient with a left frontal-insular zone 1 expansile mass who underwent intraoperative ECoG testing. The incorporation of vascular reconstructions was especially beneficial, offering precise and reliable landmarks for accurate electrode placement.

## 4. Discussion

In this study, we describe the efficacy of BrainTRACE, a novel tool for localizing ECoG electrodes in patients undergoing neurosurgical procedures requiring subdural array placement. BrainTRACE’s performance was assessed by calculating time-to-completion and measuring inter-rater reliability among a diverse group of users, including undergraduate students, research technicians, medical students, and faculty. Using intraoperative photography, users independently localized electrodes, and their placements were assessed for consistency and accuracy relative to a consensus placement. The analysis underscores BrainTRACE’s reliability, as evidenced by the high inter-rater consistency among expert users. Nevertheless, lower inter-rater consistency among novice users highlighted this task’s complex and non-trivial nature. Users should have a foundational understanding of neuroanatomy, and novice users may benefit from training in cortical anatomy prior to grid registrations, with confirmations of placements made by the implanting neurosurgeon or another experienced user.

To our knowledge, no other tools exist to enable the placement of ECoG electrodes on individual patient brain anatomy without using postoperative CT. BrainTRACE addresses a critical gap in clinical care and research applications by providing a reliable method for localizing ECoG electrodes in patients with brain tumors. Traditional localization techniques, adequate for epilepsy but insufficient for tumor-involved cases, are effectively supplemented by BrainTRACE’s innovative use of intraoperative photography and vascular mapping. More accurate data from properly localized electrodes can improve understanding of sensory and cognitive processes, along with pathological conditions, in research contexts.

The potential applications of BrainTRACE extend beyond brain cancer applications to include epilepsy and movement disorder procedures. Accurate electrode localization is crucial in epilepsy for correlating electrophysiological data with cortical regions, guiding resection, and optimizing surgical outcomes (Engel et al., 2013). Similarly, precise subdural electrode placement in deep brain stimulation (DBS) is essential for targeting functional regions and minimizing side effects (Mayberg et al., 2005). By enhancing localization accuracy, BrainTRACE contributes to improved clinical outcomes and enables more reliable mapping of cortical functions.

Despite its strengths, BrainTRACE has areas that merit further exploration. Clear vasculature extraction is beneficial to enhancing the alignment of electrode grids with cortical surfaces and improved image processing techniques are needed to reliably segment these structures. Additionally, BrainTRACE begins with the assumption that electrodes are a fixed distance (e.g., 10 mm center-to-center spacing that is common for clinical grids). While these distances are physically set at a specific distance, cortical distances may vary slightly from physical distances due to imperfections in the reconstruction of the pial surface. For example, if a high threshold is selected, the cortical surface will be eroded, and there will be less geodesic distance between the real electrode locations. Addressing these discrepancies requires refining BrainTRACE to allow within-grid refinements in spacing and accounting for brain tissue swelling and protrusion during the operation.

Furthermore, neuronavigational tools provide a potentially useful but challenging mode of grid localization. Current methods do not provide reliable or accurate coordinates. Therefore, intraoperative photography should remain the gold standard for grid placement. Future iterations of BrainTRACE could incorporate real-time ECoG grid placement and data analysis, further enhancing intraoperative decision-making. Preoperative image pre-processing would streamline integration into surgical workflows, allowing surgeons to adjust placements dynamically. BrainTRACE’s flexibility in operating with standardized brain models ensures its usability in diverse scenarios, including those with significant anatomical defects or missing imaging data. The data presented in this paper validates BrainTRACE as an essential tool for providing a reliable solution for localizing ECoG electrodes in patients with brain tumors and supporting high-quality electrophysiological data collection.

## Data Availability

All data produced in the present study are available upon reasonable request to the authors

https://github.com/dbrang/BrainTRACE

## Acknowledgments

This study was funded by NIH Grants R01DC020717 and R01NS137850.

## Declarations of Interest

Author David Brang is an inventor on a patent application (Application No. 63/785,486), which covers aspects of the software described in this study. The authors declare no other competing interests.

## Notes

### Author Declarations

The IRB of the University of California, San Francisco gave ethical approval for this work.

## References

Aabedi, A. A., Lipkin, B., Kaur, J., Kakaizada, S., Valdivia, C., Reihl, S., Young, J. S., Lee, A. T., Krishna, S., Berger, M. S., Chang, E. F., Brang, D., & Hervey-Jumper, S. L. (2021). Functional alterations in cortical processing of speech in glioma-infiltrated cortex. Proceedings of the National Academy of Sciences of the United States of America, 118(46), e2108959118. 10.1073/pnas.2108959118

Ashburner, J., & Friston, K. J. (2005). Unified segmentation. NeuroImage, 26(3), 839–851.

Avants, B. B., Tustison, N. J., Song, G., Cook, P. A., Klein, A., & Gee, J. C. (2011). A reproducible evaluation of ANTs similarity metric performance in brain image registration. Neuroimage, 54(3), 2033–2044. 10.1016/j.neuroimage.2010.09.025

Awuah, W. A., Ahluwalia, A., Darko, K., Sanker, V., Tan, J. K., Tenkorang, P. O., Ben-Jaafar, A., Ranganathan, S., Aderinto, N., Mehta, A., Shah, M. H., Lee Boon Chun, K., Abdul-Rahman, T., & Atallah, O. (2024). Bridging Minds and Machines: The Recent Advances of Brain-Computer Interfaces in Neurological and Neurosurgical Applications. World Neurosurgery, 189, 138–153. 10.1016/j.wneu.2024.05.104

Billot, B., Greve, D. N., Puonti, O., Thielscher, A., Van Leemput, K., Fischl, B., … & Iglesias, J. E. (2023). SynthSeg: Segmentation of brain MRI scans of any contrast and resolution without retraining. Medical image analysis, 86, 102789.

Branco, M. P., Leibbrand, M., Vansteensel, M. J., Freudenburg, Z. V., & Ramsey, N. F. (2018). GridLoc: An automatic and unsupervised localization method for high-density ECoG grids. NeuroImage, 179, 225–234. 10.1016/j.neuroimage.2018.06.050

Brang, D., Dai, Z., Zheng, W., & Towle, V. L. (2016). Registering imaged ECoG electrodes to human cortex: A geometry-based technique. Journal of neuroscience methods, 273, 64–73. 10.1016/j.jneumeth.2016.08.007

Chang, E. (2015). Towards Large-Scale Human-Based Mesoscale Neurotechnologies. Neuron, 86(1), 68. 10.1016/j.neuron.2015.03.037

Chang, E. F., Clark, A., Smith, J. S., Polley, M. Y., Chang, S. M., Barbaro, N. M., Parsa, A. T., McDermott, M. W., & Berger, M. S. (2011). Functional mapping-guided resection of low-grade gliomas in eloquent areas of the brain: improvement of long-term survival. Journal of Neurosurgery, 114(3), 566–573. 10.3171/2010.6.JNS091246

Crone, N. E., Boatman, D., Gordon, B., & Hao, L. (2001). Induced electrocorticographic gamma activity during auditory perception. Clinical Neurophysiology, 112(4), 565–582. 10.1016/s1388-2457(00)00545-9

Dalal, S. S., Edwards, E., Kirsch, H. E., Barbaro, N. M., Knight, R. T., & Nagarajan, S. S. (2008). Localization of neurosurgically implanted electrodes via photograph-MRI-radiograph coregistration. Journal of Neuroscience Methods, 174(1), 106–115. 10.1016/j.jneumeth.2008.06.028

Engel, A. K., Moll, C. K., Fried, I., & Ojemann, G. A. (2005). Invasive recordings from the human brain: Clinical insights and beyond. Nature Reviews Neuroscience, 6(1), 35–47. 10.1038/nrn1585

Fischl, B. (2012). FreeSurfer. NeuroImage, 62(2), 774–781.

Ganesan, K., Plass, J., Beltz, A. M., Liu, Z., Grabowecky, M., Suzuki, S., Stacey, W. C., Wasade, V. S., Towle, V. L., Tao, J. X., Wu, S., Issa, N. P., & Brang, D. (2021). Visual speech differentially modulates beta, theta, and high gamma bands in auditory cortex. The European Journal of Neuroscience, 54(9), 7301. 10.1111/ejn.15482

Ghuman, A. S., Brunet, N. M., Li, Y., Konecky, R. O., Pyles, J. A., Walls, S. A., Destefino, V., Wang, W., & Richardson, R. M. (2014). Dynamic encoding of face information in the human fusiform gyrus. Nature Communications, 5(1), 1–10. 10.1038/ncomms6672

Gupta, D., Hill, N. J., Adamo, M. A., Ritaccio, A., & Schalk, G. (2014). Localizing ECoG electrodes on the cortical anatomy without post-implantation imaging. NeuroImage: Clinical, 6, 64. 10.1016/j.nicl.2014.07.015

Hervey-Jumper, S. L., Li, J., Lau, D., Molinaro, A. M., Perry, D. W., Meng, L., & Berger, M. S. (2015). Awake craniotomy to maximize glioma resection: methods and technical nuances over a 27-year period. Journal of Neurosurgery, 123(2), 325–339. 10.3171/2014.10.JNS141520

Kingyon, J., Behroozmand, R., Kelley, R., Oya, H., Kawasaki, H., Narayanan, N. S., & W Greenlee, J. D. (2015). High-gamma band fronto-temporal coherence as a measure of functional connectivity in speech motor control. Neuroscience, 305, 15. 10.1016/j.neuroscience.2015.07.069

Klaes, C. (2018). Chapter 28 - Invasive Brain-Computer Interfaces and Neural Recordings From Humans. In D. Manahan-Vaughan (Ed.), Handbook of Behavioral Neuroscience (Vol. 28, pp. 527–539). Elsevier. 10.1016/B978-0-12-812028-6.00028-8

Krishna, S., Choudhury, A., Keough, M. B., Seo, K., Ni, L., Kakaizada, S., Lee, A., Aabedi, A., Popova, G., Lipkin, B., Cao, C., Nava Gonzales, C., Sudharshan, R., Egladyous, A., Almeida, N., Zhang, Y., Molinaro, A. M., Venkatesh, H. S., Daniel, A. G., … L., S. (2023). Glioblastoma remodelling of human neural circuits decreases survival. Nature, 617(7961), 599–607. 10.1038/s41586-023-06036-1

Merk, T., Peterson, V., Lipski, W. J., Blankertz, B., Turner, R. S., Li, N., Horn, A., Richardson, R. M., & Neumann, W.-J. (2022). Electrocorticography is superior to subthalamic local field potentials for movement decoding in Parkinson’s disease. eLife, 11:e75126

Moon, H., Kwon, J., Eun, J., Chung, C. K., Kim, J. S., Chou, N., & Kim, S. (2024). Electrocorticogram (ECoG): Engineering Approaches and Clinical Challenges for Translational Medicine. Advanced Materials Technologies, 9(12), 2301692. 10.1002/admt.202301692

Pieters, T. A., Conner, C. R., & Tandon, N. (2013). Recursive grid partitioning on a cortical surface model: an optimized technique for the localization of implanted subdural electrodes. Journal of Neurosurgery, 118(5), 1086–1097. 10.3171/2013.2.JNS121450

Ray, S., Crone, N. E., Niebur, E., Franaszczuk, P. J., & Hsiao, S. S. (2008). Neural correlates of high-gamma oscillations (60–200 Hz) in macaque local field potentials and their potential implications in electrocorticography. Journal of Neuroscience, 28(45), 11526–11536.

Sacino, M. F., Ho, C. Y., Murnick, J., Tsuchida, T., Magge, S. N., Keating, R. F., Gaillard, W. D., & Oluigbo, C. O. (2016). Intraoperative MRI-guided resection of focal cortical dysplasia in pediatric patients: technique and outcomes. Journal of Neurosurgery: Pediatrics, 17(6), 672–678. 10.3171/2015.10.PEDS15512

Spencer, S. S., Spencer, D. D., Williamson, P. D., & Mattson, R. (1990). Combined depth and subdural electrode investigation in uncontrolled epilepsy. Neurology, 40(1), 74–79. 10.1212/wnl.40.1.74

Wellmer, J., von Oertzen, J., Schaller, C., Urbach, H., König, R., Widman, G., Van Roost, D., & Elger, C. E. (2002). Digital photography and 3D MRI-based multimodal imaging for individualized planning of resective neocortical epilepsy surgery. Epilepsia, 43(12), 1543–1550. 10.1046/j.1528-1157.2002.30002.x

